# Association between *CCR5*-Δ32 homozygosity and mortality in 37,650 participants from three U.S.-based cohorts

**DOI:** 10.1101/19006619

**Authors:** Xia Jiang, Hongyan Huang, Francine Grodstein, Peter Kraft

## Abstract

An analysis of 409,693 UK Biobank participants recently published in *Nature Medicine* identified a relative 21% increase in all-cause mortality among participants who were homozygous for the Δ32 deletion in the C-C motif chemokine receptor 5 gene (*CCR5*).^1^ This is a timely and potentially cautionary result in light of He Jiankui’s controversial germline editing of *CCR5* to induce mutations that putatively mimic the effects of Δ32, which is known to reduce the risk of HIV infection. To provide additional evidence on the association between the Δ32 deletion and mortality and assess its generalizability, we present results from three large-scale population-based US cohorts: the Nurses’ Health Study (NHS),^2^ the NHSII and the Health Professional Follow-Up Study (HPFS).^3^

---

We followed 37,650 participants (NHS 18,496; NHSII 8,276; HPFS 10,878) who donated blood samples *circa* 1990 (NHS), 1994 (HPFS) and 1996 (NHS2). These participants were genotyped as previous genome-wide association studies of several chronic diseases (including breast and colon cancers and Type 2 Diabetes); all participants were disease-free at blood draw. Two variants (rs113341849 and rs113010081), were used as proxies for *CCR5*-Δ32 (rs333) (*r*^2^=0.97 and 0.94 in European 1,000 Genomes Project samples, respectively). Participant deaths were identified via multiple sources, with documented 98% accuracy through July 2019. We used Cox proportional hazards regression to estimate hazard ratios (HR) of death in relation to *CCR5*-Δ32 homozygosity (Δ32/Δ32 *vs*. Δ32/+ or +/+), counting person-time from age at blood draw to death or end of study period, whichever occurred first.

During a median 21.8 years of follow-up, we documented 12,530 deaths (NHS 7,146; NHSII 306; HPFS 5,078). In contrast to Wei *et al*.’s report with 13,831 deaths, neither SNP deviated from Hardy-Weinberg equilibrium in our samples at blood draw: the ratio of observed to expected heterozygotes was 1.00 for both rs113341849 and rs113010081 (P=0.64 and P=0.85). We did not find strong support for heightened mortality among *CCR5*-Δ32 homozygotes in our pooled cohorts (rs113341849: HR=1.08 [95%CI: 0.89-1.32], P=0.44; rs113010081: 1.05 [0.85-1.29], P=0.68). Analysis within the all-female NHS and NHS2 cohorts provided little evidence for association with mortality (rs113341849: 0.99 [0.76-1.30], P=0.95; rs113010081: 0.94 [0.70-1.25], P=0.66). For the all-male HPFS, we observed an apparent 20-22% increased mortality rate, although with substantial statistical uncertainty (rs113341849: 1.22 [0.91-1.65], P=0.19; rs113010081: 1.20 [0.89-1.63], P=0.24). There was little evidence for differences in mortality associations between females and males (heterogeneity P=0.31 and P=0.25).

Overall, we did not find compelling evidence for a link between *CCR5* deletion and shortened lifespan. Our cohorts and UK Biobank share similarities, including European-ancestry participants, prevalence of exposure (*CCR5*-Δ32 allele frequency ∼10%) and comparable number of deaths. Several differences may explain discrepancies in results. While Wei *et al*. used directly genotyped markers as proxies to identify *CCR5*-Δ32, we used imputed markers (imputation *r*^2^=0.69-0.84). Although disease-free at blood draw, our subjects are an ascertained sample who lived long enough to be diagnosed with a disease or selected as an age-matched control. Still, most deaths occurred after the ages at which our target diseases were diagnosed (75% of deaths in these cohorts occurred after age 76), and the distribution of age at death in our analytic sample is similar to that in the underlying full cohort; this minimizes the likelihood and magnitude of bias due to ascertainment. In conclusion, we do not find strong evidence supporting an association between *CCR5*-Δ32 homozygosity and mortality in a large sample from three US-based cohorts, suggesting that if present, the mortality association is weaker than estimated from the original report.

**Fig1.**
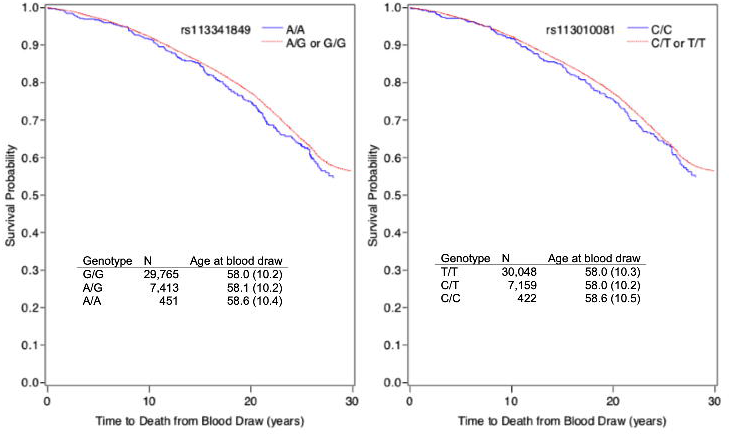
Kaplan-Meier survival curve for rs113341849 (left panel) and rs113010081 (right panel). X-axis indicates time from blood draw; y-axis indicates survival probability. The rs113341849 A and rs113010081 C alleles are proxies for *CCR5*-Δ32. Age: mean (standard deviation) age at blood draw.

## Data Availability

Data are available upon request, see: https://www.nurseshealthstudy.org/researchers and https://sites.sph.harvard.edu/hpfs/for-collaborators/.

## Acknowledgements

The Institutional Review Boards at The Brigham and Women’s Hospital and Harvard T.H. Chan School of Public Health IRBs approved the study protocol. All subjects provided informed consent. This work was supported by U.S. National Institutes of Health Grants U01 CA186107, R01 CA49449, U01 CA176726, R01 CA67262, and U01 CA167552. Dr. Jiang is supported by the International Postdoc Grant from the Swedish Research Council. Data are available upon request, see: https://www.nurseshealthstudy.org/researchers and https://sites.sph.harvard.edu/hpfs/for-collaborators/. Code is available upon request from the corresponding author.

## References

1. Wei, X. & Nielsen, R. CCR5-∆32 is deleterious in the homozygous state in humans. Nat. Med. 25, 909–910 (2019).

2. Colditz, G. A., Manson, J. E. & Hankinson, S. E. The Nurses’ Health Study: 20-year contribution to the understanding of health among women. J. Womens Health 6, 49–62 (1997).

3. Rimm, E. B., Stampfer, M. J., Colditz, G. A., Giovannucci, E. & Willett, W. C. Effectiveness of various mailing strategies among nonrespondents in a prospective cohort study. Am. J. Epidemiol. 131, 1068–1071 (1990).

